# The implementation of KPI (key performance indicators) to manage the Pharmacovigilance System of a public vaccine producer in Brazil

**DOI:** 10.1101/2025.02.23.25322752

**Authors:** Mayra M. M. Oliveira, Fernanda L. C. Oliveira, Vera L. Gattás, Clarissa L. Santos, Gabriela A. Wagner, Monica Taminato

**Affiliations:** Instituto Butantan, Clinical Trials and Pharmacovigilance Division, São Paulo, Brazil; Federal University of São Paulo, São Paulo, Brazil

**Author notes:** Corresponding author (MMMO).

**Keywords:** Pharmacovigilance, Key Performance Indicators (KPIs), Quality Improvement

## Abstract

**Background:** Pharmacovigilance (PV) is a critical science aimed at enhancing patient safety. For a pharmacovigilance system (PVS) to be effective, it must be robust, efficient, and regularly evaluated to ensure ongoing improvement. Key Performance Indicators (KPIs) are essential quantifiable measures that evaluate the performance of companies, processes, and systems, including pharmacovigilance.

**Aim:** This study aimed to implement and evaluate KPIs for the PVS of a public vaccine producer in Brazil, identifying improvement areas and supporting continuous enhancement.

**Methods:** KPI selection for Instituto Butantan’s PVS was guided by a systematic review and benchmarking with pharmaceutical representatives, followed by validation through expert consensus. A dashboard was created for KPI implementation, with goals, measurements, and evaluation frequency detailed in the institution’s Standard Operating Procedure (SOP).

**Results:** The review identified one KPI for a Marketing Authorization Holder (MAH) PV process. Interviews with pharmaceutical company representatives led to 25 additional KPIs, with one overlapping. After expert review, all KPIs were approved after adjustments, and 10 were piloted, including metrics like the number of individual case safety reports (ICSR), ICSR per product, and serious adverse events (SAE) per product. A dashboard tracked these KPIs monthly in 2022 and 2023.

**Conclusion:** Over two years, KPI evaluations proved valuable, prompting improvements in PV processes and tools. Results indicated that expanding to the remaining KPIs would further enhance Instituto Butantan’s PVS.

Pharmacovigilance, Key Performance Indicators (KPIs), Quality Improvement

## Introduction

Pharmacovigilance (PV) is the science whose activities include detecting, assessing, understanding, and preventing adverse events and other problems related to medications. A Pharmacovigilance System (PVS) is designed to monitor the safety of medications and detect any changes in their risk-benefit relationship. Therefore, the PVS must be robust and efficient enough to produce relevant results on time for necessary actions and needs to be continuously evaluated for its quality [1].

The quality of a PVS can be measured by predefined requirements, which are the system’s characteristics likely to produce the desired outcome or quality objectives. General quality objectives for a PVS include [2]:

- Meeting legal requirements for PV tasks and responsibilities;
- Preventing harm caused by adverse reactions in humans from the use of medications, whether within or outside the terms of market authorization or occupational exposure;
- Promoting the safe and effective use of medications, particularly through providing safety information to patients, healthcare professionals, and the public; and
- Contributing to the protection of patients and public health.

Key performance indicators (KPIs) are quantitative measures used to assess the quality of critical processes within a company or department, such as PV. By interpreting KPI results, measures for the continuous improvement of a marketing authorization holder’s (MAH) PVS can be identified and adopted to make it more robust and ensure that quality objectives are met [2]. Instituto Butantan (IB) is a Brazilian public institution and is the largest producer of immunobiologicals, serums, and vaccines in Brazil [3]. As a MAH, IB has been monitoring the safety of its products through a consolidated PVS since 2010. In 2017, IB received consultancy from PATH, a global non-profit health organization, to strengthen its PVS, with one of the identified needs being the implementation of KPIs [4].

Therefore, in this study we aimed to implement and evaluate KPIs for the PVS of Instituto Butantan, a public Brazilian vaccine MAH. to identify areas for improvement, maintain system efficiency, and support continuous enhancement.

## Materials and methods

To define the KPIs, a systematic review, benchmarking with 5 companies, and a validation process through expert consensus were conducted.

### Systematic Review

The systematic review followed the steps proposed by the Cochrane Collaboration. Studies were included regardless of language or publication format. Inclusion criteria required that studies address PV and some KPIs.

Scientific evidence was selected through an electronic search of the Cochrane Library database (including the Cochrane Controlled Trials Register), PUBMED, EMBASE, Web of Science, LILACS, SciELO, abstracts from conference presentations, and references from published review and systematic review articles. The search strategy used is outlined in Fig1 for Pubmed and adapted for other databases. Publications were managed in Mendeley to remove duplicates and apply the inclusion criteria.

**Fig 1.**
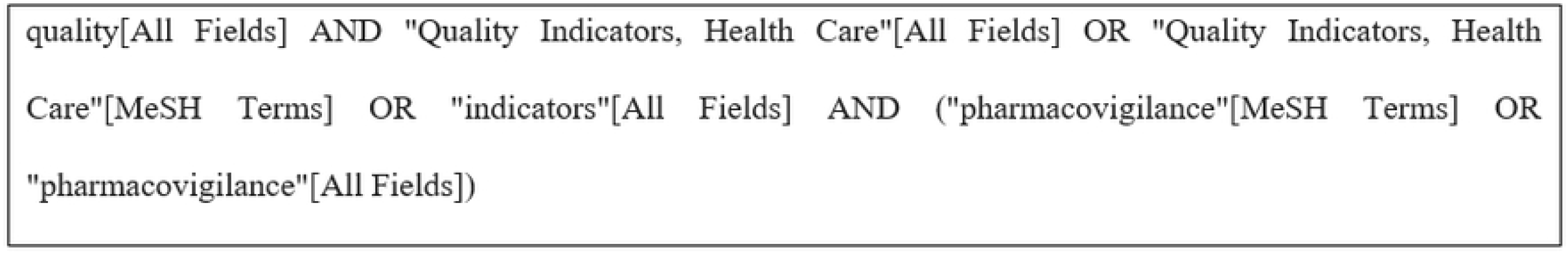
Search Strategy for PubMed.

### Benchmarking

A research instrument was developed to identify which KPIs are used and how they are utilized by other national and multinational pharmaceutical companies that produce or distribute immunobiological in Brazil.

Currently, in Brazil, there are three public companies, in addition to Instituto Butantan, that produce or distribute immunobiologicals, and six private companies operating in the field. The research instrument was administered remotely by the project’s researcher via videoconferences. An invitation was sent to the Qualified Person Responsible for Pharmacovigilance (QPPV) of each company electronically. The QPPV could either be the interviewee or designate another professional from the PV department.

All interviewees signed the Informed Consent Form (ICF), and once it was signed, the interview commenced using the research instrument. The purpose of the instrument was to assess which indicators are used by these PV departments, how these indicators are monitored and updated, and to understand the existing procedures for utilizing them.

### Validation of the indicators through expert consensus

The validation of the KPIs was conducted through a consensus meeting via video conference using a methodology adapted Delphi [5] employing the Nominal Group Technique (NGT) [6]. Seven participants were invited.

The Expert Consensus panel consisted of 6 professionals representing: the pharmaceutical industry in Brazil, medical societies, scientific academia, Brazil’s representatives in the Vaccine Safety Net of the World Health Organization, and representatives from the Epidemiological Surveillance of the State of São Paulo, Brazil. No representatives from the companies interviewed during the benchmarking process were invited to participate as experts in the KPI validation process.

All experts attended a lecture to understand what a PVS is and the purpose of the selected KPIs. Each KPI was presented in a technical sheet format that included: 1) Indicator name, 2) Indicator type, 3) Definition, Formula for calculating the indicator, 5) Evaluation frequency of the indicator, 6) Data source, 7) Observations with references and definitions of technical terms.

Participants were invited and given basic instructions about the workshop, including the date and time. They also received in advance the materials containing instructions on how to participate in the consensus process, detailed technical sheets for the 25 KPIs previously defined after the literature review and benchmarking along with some references [7-11].

The experts were able to watch a recorded lecture explaining the purpose of these KPIs before evaluating each technical sheet and scoring them using RedCap, rating from 1 to 9 whether the KPI was Inadequate, Undefined, or Adequate. KPIs were classified into three levels based on the calculated median, referred to as the “three-point region”. Those with a median between 1-3 were considered inadequate, 4-6 were deemed undefined, and 7-9 were considered adequate.

KPIs with a median of 3 or less were excluded, while those with a median of 4 to 6 were sent for a second round of consensus. An Informed Consent Form (ICF) was presented and signed by each participant, containing information about their rights and assurances of confidentiality and privacy. All experts signed the ICF before accessing the materials and performing their evaluations.

The ChatGPT language model, developed by OpenAI, was used to enhance the clarity and cohesion of the text in this article.

## Results

The KPIs defined by the previously described methods were validated through a consensus of six experts. To implement the indicators, a dashboard was created, and the KPIs, along with their goals, measurements, and evaluation frequency, were detailed in the Instituto Butantan’s Standard Operating Procedure (SOP).

### Key Performance Indicators identified by the systematic review

Our results show that approximately 50% of the 25 studies included in this scoping review were conducted in the past 8 years, indicating a recent focus on the quality management of PVS. A total of 87 PV indicators were identified, of which 7 (8.0%) pertain to signal detection, and only one is related to a PV KPI for MAHs (Table 1) [12].

**Table 1.**
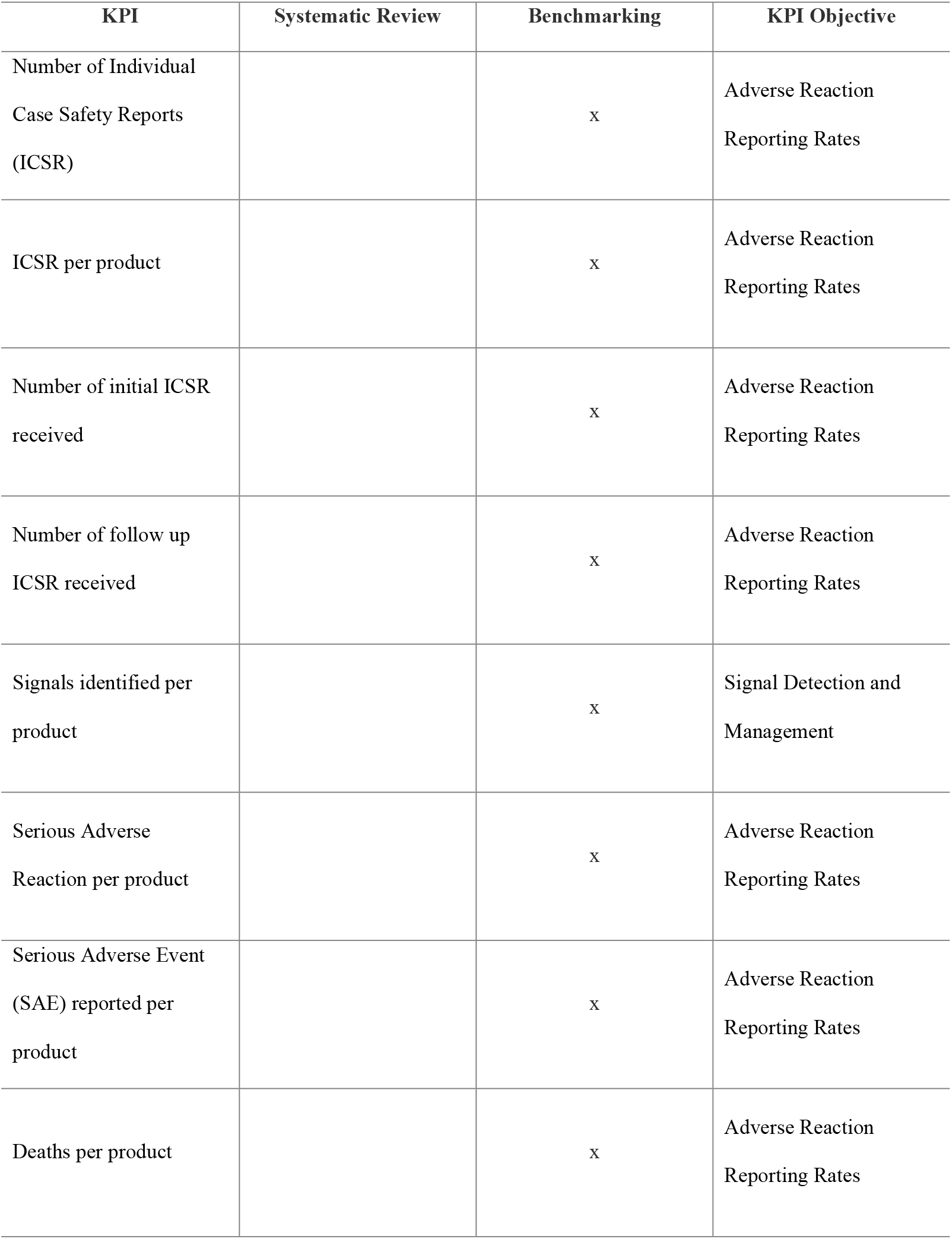
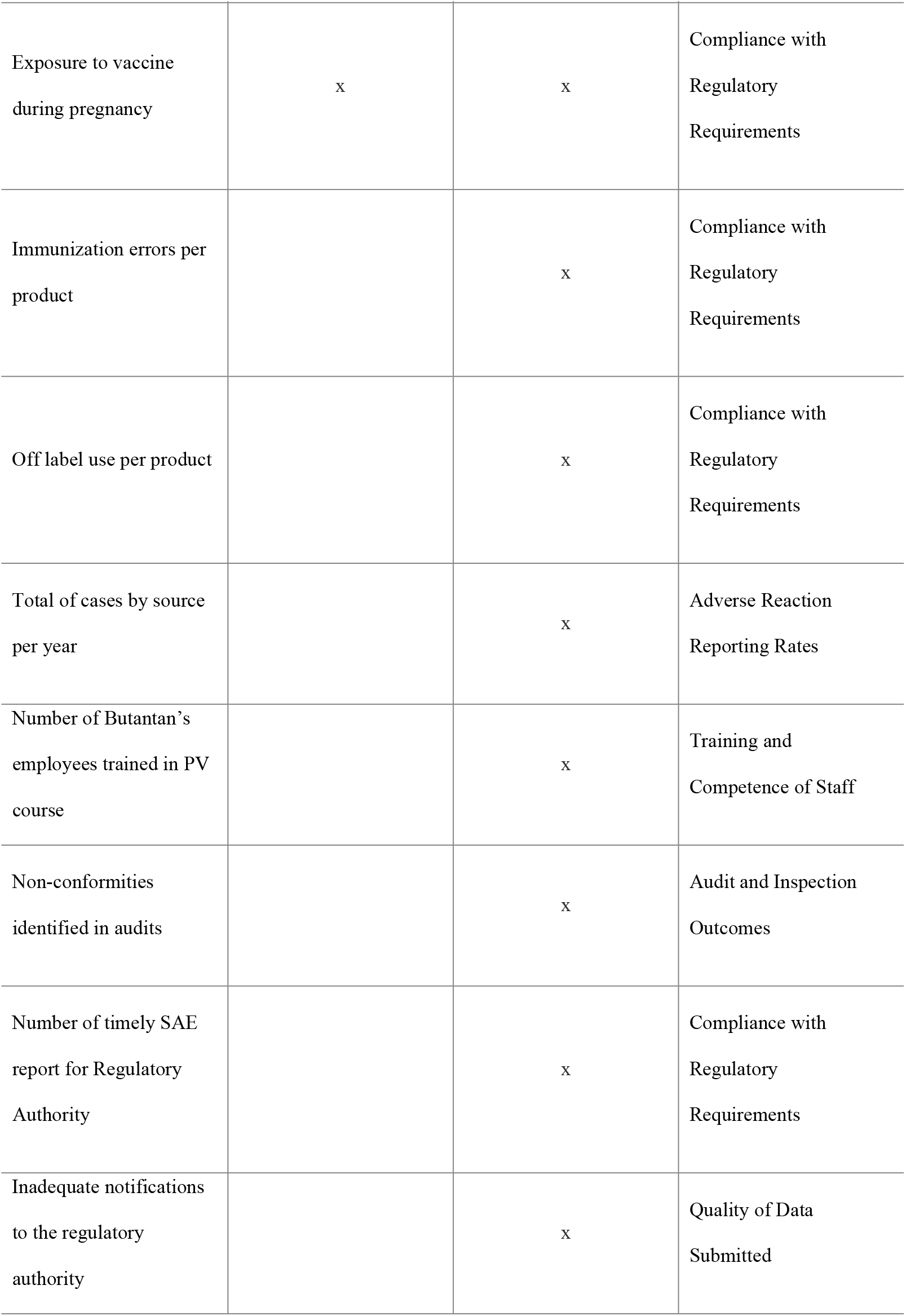
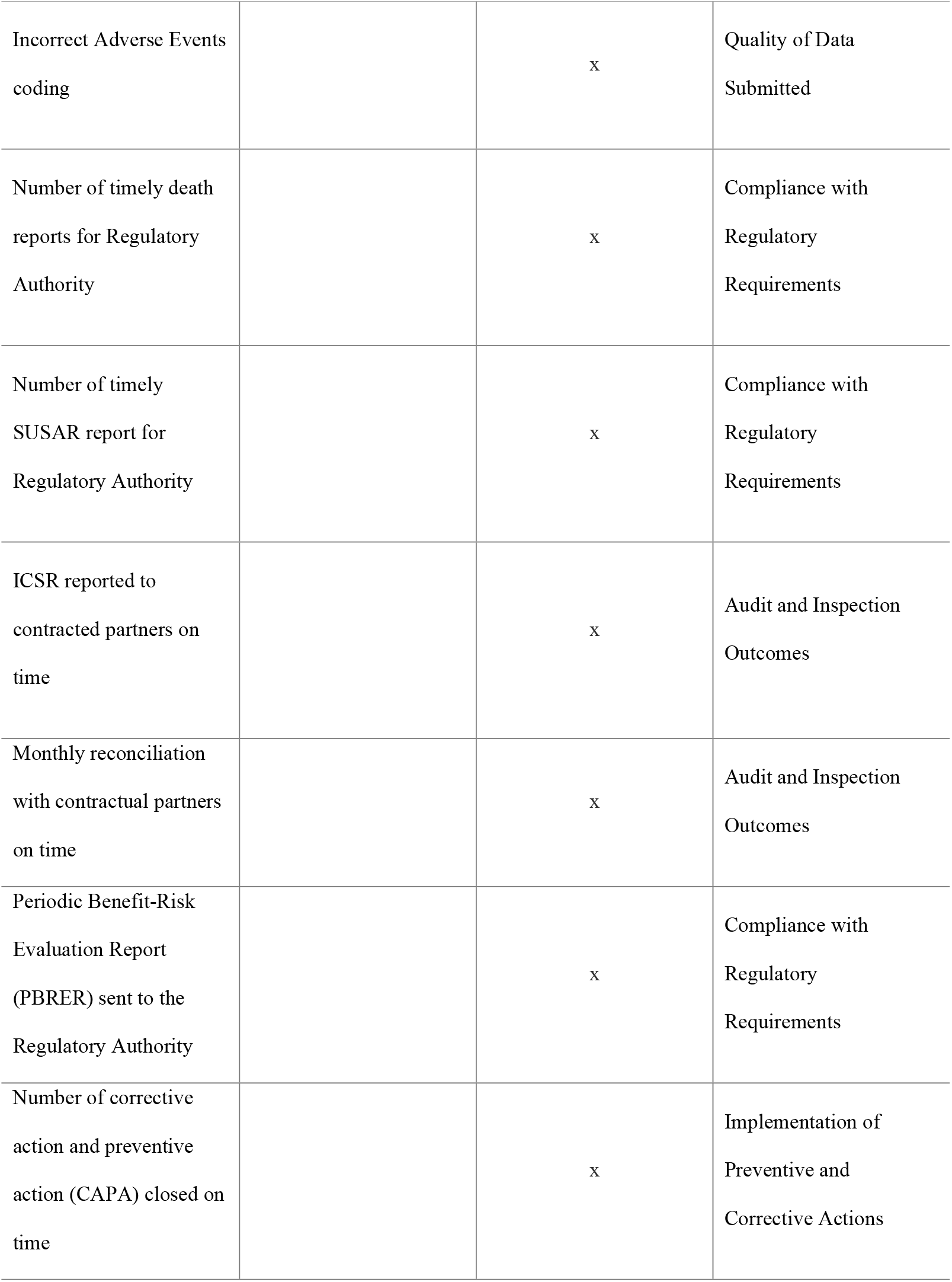
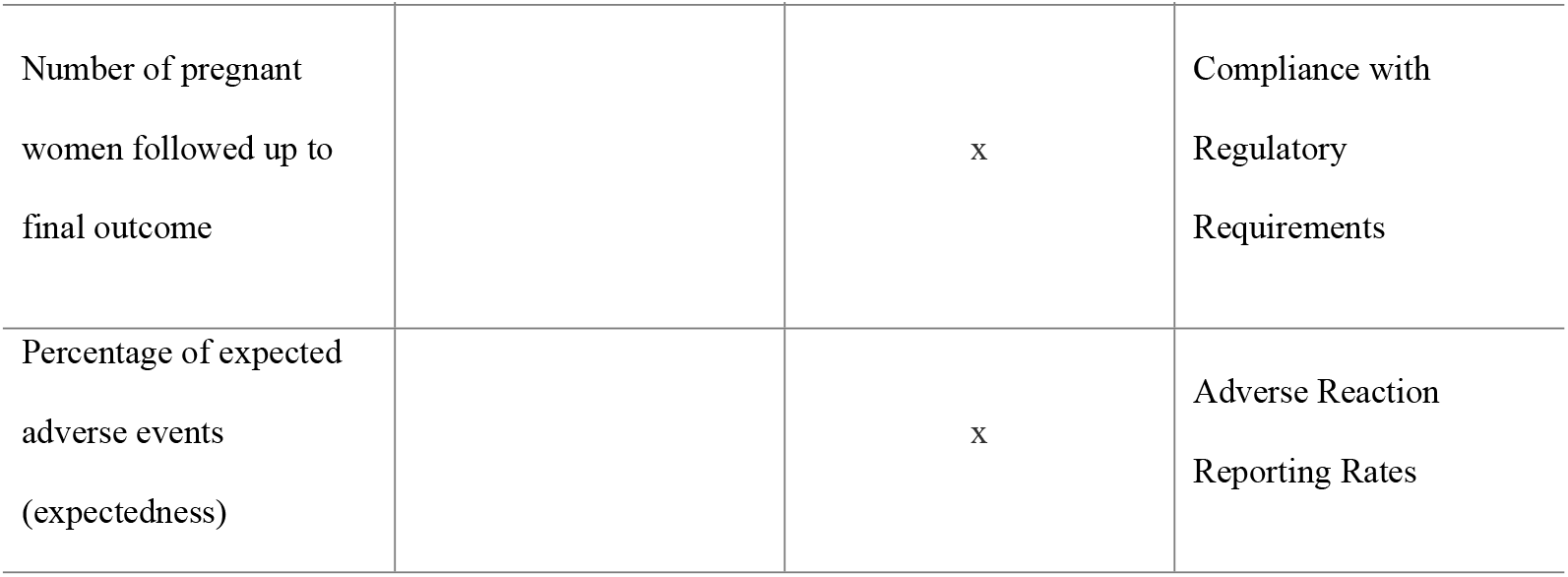
KPIs identified by benchmarking and systematic review and their objectives.

| KPI | Systematic Review | Benchmarking | KPI Objective |
| --- | --- | --- | --- |
| Number of Individual Case Safety Reports (ICSR) |  | x | Adverse Reaction Reporting Rates |
| ICSR per product |  | x | Adverse Reaction Reporting Rates |
| Number of initial ICSR received |  | x | Adverse Reaction Reporting Rates |
| Number of follow up ICSR received |  | x | Adverse Reaction Reporting Rates |
| Signals identified per product |  | x | Signal Detection and Management |
| Serious Adverse Reaction per product |  | x | Adverse Reaction Reporting Rates |
| Serious Adverse Event (SAE) reported per product |  | x | Adverse Reaction Reporting Rates |
| Deaths per product |  | x | Adverse Reaction Reporting Rates |

| **KPI** | **Systematic Review** | **Benchmarking** | **KPI Objective** |
| --- | --- | --- | --- |
| Exposure to vaccine during pregnancy | x | x | Compliance with Regulatory Requirements |
| Immunization errors per product |  | x | Compliance with Regulatory Requirements |
| Off label use per product |  | x | Compliance with Regulatory Requirements |
| Total of cases by source per year |  | x | Adverse Reaction Reporting Rates |
| Number of Butantan's employees trained in PV course |  | x | Training and Competence of Staff |
| Non-conformities identified in audits |  | x | Audit and Inspection Outcomes |
| Number of timely SAE report for Regulatory Authority |  | x | Compliance with Regulatory Requirements |
| Inadequate notifications to the regulatory authority |  | x | Quality of Data Submitted |
| Incorrect Adverse Events coding |  | x | Quality of Data Submitted |
| Number of timely death reports for Regulatory Authority |  | x | Compliance with Regulatory Requirements |
| Number of timely SUSAR report for Regulatory Authority |  | x | Compliance with Regulatory Requirements |
| ICSR reported to contracted partners on time |  | x | Audit and Inspection Outcomes |
| Monthly reconciliation with contractual partners on time |  | x | Audit and Inspection Outcomes |
| Periodic Benefit-Risk Evaluation Report (PBRER) sent to the Regulatory Authority |  | x | Compliance with Regulatory Requirements |
| Number of corrective action and preventive action (CAPA) closed on time |  | x | Implementation of Preventive and Corrective Actions |
| Number of pregnant women followed up to final outcome |  | x | Compliance with Regulatory Requirements |
| Percentage of expected adverse events (expectedness) |  | x | Adverse Reaction Reporting Rates |

### KPIs identified by benchmarking

As a result of the benchmarking, 25 indicators were obtained, of which only 1 had also been mentioned in the literature review results (Table 1).

### Validation of KPIs by expert consensus

For the validation, seven experts were invited, however the representative from Anvisa (Brazilian Regulatory Agency) was unable to attend. No KPIs were deemed inadequate by expert consensus. Some received median scores between 7 and 9, which, according to the methodology, would have been sufficient for approval without needing a consensus meeting. However, due to lower scores from some experts, these KPIs were also discussed in the meeting.

During the meeting, experts highlighted the need to improve the definitions of the KPIs to clarify their purposes and suggested revisiting the calculations of some indicators. Ten KPIs were selected for pilot implementation in the PVS of the Instituto Butantan, designated as the first implementation block (Table 2).

**Table 2.** KPIs designated as the first implementation block.

| Number of KPI | Name of KPI before validation | Name of KPY after validation |
| --- | --- | --- |
| 1 | Number of individual case safety reports (ICSR) | Total cases processed by the PV team in the period |
| 2 | ICSR per product | Cases by product in the period |
| 3 | Serious Adverse Event (SAE) reported per product | Serious and non-serious cases in the period |
| 4 | Exposure to vaccine during pregnancy | Cases of pregnant women by product in the period |
| 5 | Number of Butantan's employees trained in PV course | IB employees' adherence to PV training |
| 6 | Number of timely SAE report for Regulatory Authority | SAE Reports notified on time to the Regulatory Authority |
| 7 | Number of timely death report for Regulatory Authority | Death reports notified on time to the Regulatory Authority |
| 8 | Number of timely SUSAR report for Regulatory Authority | SUSAR notified on time to the Regulatory Authority |
| 9 | ICSR reported to contracted partners on time | Reports exchanged with Contractual Partner on time |
| 10 | Number of corrective action and preventive action (CAPA) closed on time | Number of corrective action and preventive action (CAPA) closed on time |

### Implementation of KPIs designated as the first block

After validating and selecting 10 KPIs for the pilot implementation of the first block, a Standard Operating Procedure (SOP) was developed. The SOP outlines each KPI’s name, definition, source, calculation method, goal, and evaluation frequency. Each month, the KPIs are evaluated and presented to the QPPV during a meeting. A dashboard was created to analyze and monitor the KPIs (Fig 2).

**Fig 2.**
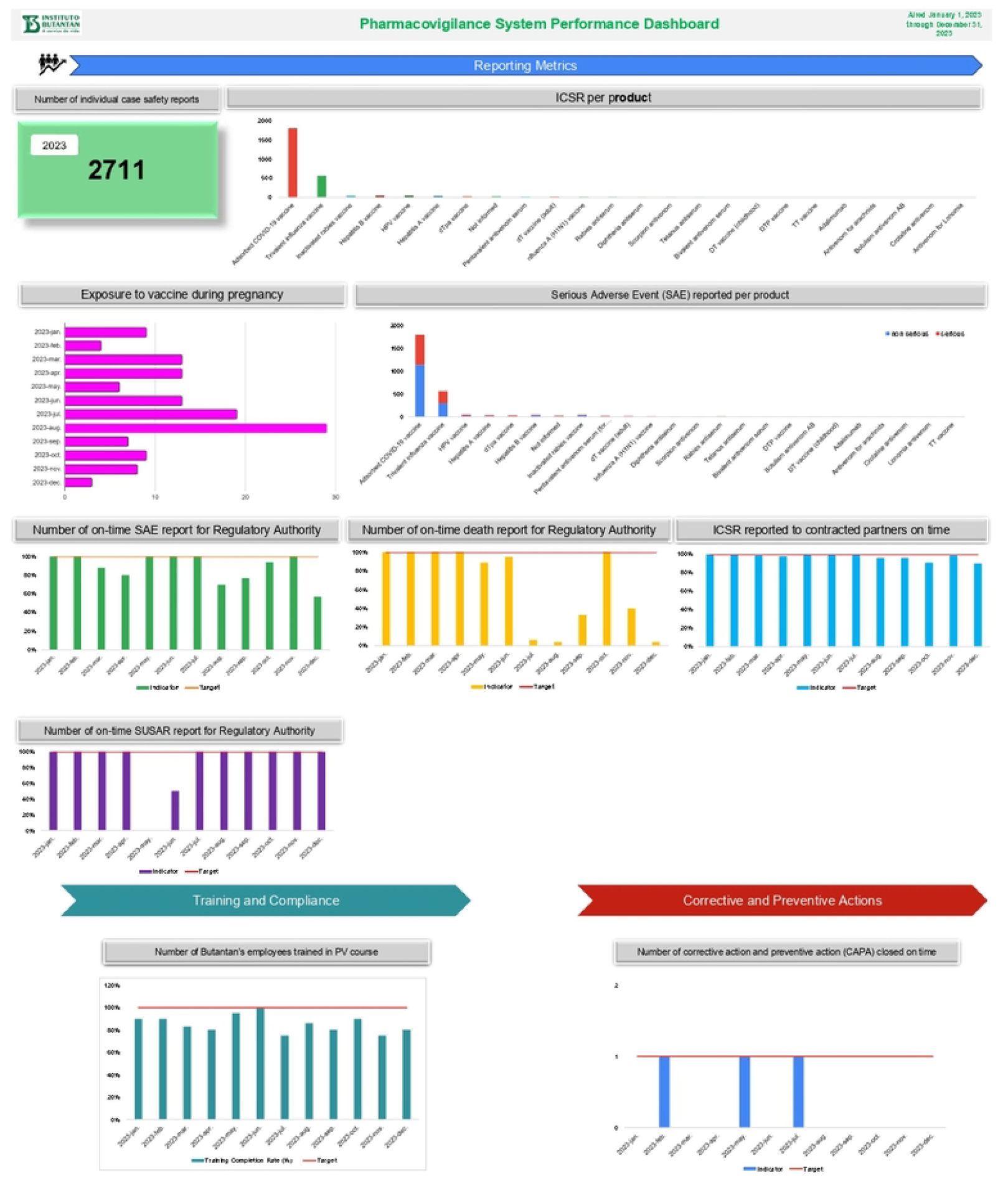
Representation of the dashboard with the 10 indicators selected for the pilot project. ICSR: individual case safety report. HPV: human papilloma virus. dT: diphtheria and tetanus. DT: diphtheria and tetanus (pediatrics). DTP: diphtheria, tetanus, and pertussis. TT: tetanus toxin. SUSAR: suspected unexpected serious adverse reaction.

The 10 selected KPIs were monitored monthly through a dashboard during 2022 and 2023 as part of a pilot to assess the significance of measuring PV processes and their impact on the PVS. Supporting information SI1 presents the 10 indicators with their objectives, frequency, targets, and calculation formulas, if applicable.

## Discussion

This manuscript describes the 25 KPIs identified through systematic review and benchmarking, validated by expert consensus, and details the implementation of 10 KPIs designated as the first implementation block for measuring, assessing, and managing the PVS at Instituto Butantan, a public Brazilian vaccine MAH.

The MAH is responsible for continuously monitoring the safety of its medicinal products, informing authorities of any changes that might impact the marketing authorization, and ensuring that product information remains up-to-date [8,10,11].

A PVS is defined as a framework used by an organization to fulfill its legal tasks and responsibilities related to PV. It is designed to monitor drug safety and detect any changes in the risk-benefit balance [2]. Quality can be measured. To determine if the required quality level has been achieved, predefined quality requirements are necessary. The organization may use performance indicators to continuously monitor the effectiveness of PV activities concerning these quality requirements [2].

In our study, we identified 25 KPIs to be implemented for measuring and monitoring the Instituto Butantan’s PVS (Table 1). The experts who evaluated these KPIs deemed all 25 to be important. However, since Instituto Butantan did not have any KPIs or established procedures before this study, the experts recommended implementing the KPIs in two phases. Consequently, 10 KPIs were selected for the first phase, which is designated as a pilot.

Organizations are encouraged to develop a systematic approach for creating and monitoring KPIs, which should align with risk-based priorities focusing on patient safety and regulatory compliance. This includes establishing a Standard Operating Procedure (SOP) for KPI management and ensuring that metrics are regularly reviewed and updated as needed to assess their relevance, and targets can be adjusted as necessary. Monitoring PVS performance allows for the implementation of corrective and preventive measures, leading to ongoing improvements. Establishing a KPI system enables PV organizations to assess the health of their systems to ensure they are functioning effectively and efficiently, verify compliance, oversee delegated responsibilities from vendors and partners, and prepare for audits and inspections [7, 13, 14].

An SOP was created to define the processes for monitoring the KPIs. It details the 10 KPIs selected for the pilot implementation, including their definitions, sources, calculation methods, target values, and evaluation frequency. Each KPI target is categorized as follows: green (target achieved), yellow (target not achieved but close), and red (target not achieved and significantly off).

The SOP also outlines how these KPIs will be presented to the QPPV and manager team. Each month, the KPIs are reviewed during a meeting with the QPPV and manager team, and the presentation includes the values for each KPI, whether targets were met, and explanations for any unmet targets or significant changes in values. The meeting includes a discussion about possible actions to be taken to enhance the results, like team training and procedure improvements. Furthermore, the SOP specifies that if a KPI target is not met for three consecutive months, a deviation must be recorded to investigate the root cause of the issue and implement a CAPA.

To effectively manage this responsibility, the KPIs might include [2, 7, 13, 15]: Adverse Reaction Reporting Rates, Compliance with Regulatory Requirements, Quality of Data Submitted, Signal Detection and Management, Training and Competence of Staff, Audit and Inspection Outcomes, Implementation of Preventive and Corrective Actions.

All 25 KPIs meet the objectives outlined above (Table 1). However, of the 10 KPIs selected for the pilot, one indicator intended for Signal Detection and Management was not included. This decision was due to challenges with the information source, as this process undergoes frequent changes that affect how data is collected. For the pilot phase, we opted to implement KPIs whose processes and information sources were more established, avoiding significant changes that could impact the development of the KPI monitoring dashboard.

A limitation of our study is the restricted number of references available on effective indicators for managing a PVS for a MAH. Our systematic review identified only 4 studies focusing on indicators for MAHs, all centered on signal detection. Consequently, we had to resort to less reliable sources, such as websites of consulting companies specializing in this area. Another challenge in conducting this study was that the team dedicated to this evaluation and implementation is very small. The entire process of assessing the best indicators for implementation, developing the SOP, and building the dashboard was carried out by the team while they were also handling other routine activities. As a result, the implementation was slower than we liked.

This study contributed to a better understanding of what should be the KPIs to be used in a PVS of a MAH and shows the process to implement those KPIs in a MAH. We will continue with the implementation of the indicators designated for block 2. It will also focus on improving processes and conducting ongoing evaluations to consider the inclusion of additional indicators not covered in this study.

## Conclusion

In this study, we identified the KPIs to measure, assess, and manage the PVS at Instituto Butantan, a public Brazilian vaccine MAH. The KPIs defined by systematic review and benchmarking were validated through a consensus of six experts. To implement the indicators, a dashboard was created, and the KPIs, along with their goals, measurements, and evaluation frequency, were detailed in the Instituto Butantan’s Standard Operating Procedure (SOP). The 10 indicators were assessed during 2022 and 2023. After these two years of implementation and evaluation, the importance of measuring the processes carried out by the PVS at Instituto Butantan and the impact of this activity on the PVS was demonstrated. Through regular evaluations, some improvements were incorporated into the routine, and even the tools used in the PVS were enhanced. Based on the pilot results, the feasibility of implementing additional KPIs is both possible and important for the continuous improvement of the PVS.

## Data Availability

All relevant data are within the manuscript and its Supporting Information files.

## Acknowledgements

With acknowledgment and thanks to the Instituto Butantan and the Paulista School of Nursing of the Federal University of São Paulo.

## Supporting information

**SI1. Key Performance Indicators – objectives, frequency, targets, and formulas**

